# Persistent Iatrogenic Atrial Septal Defect Following Pulsed Field Ablation Guided by Left Atrial Intracardiac Echocardiography: Incidence, Predictors, and Clinical Outcomes

**DOI:** 10.64898/2026.06.30.26356981

**Authors:** Mohamed Mraiyan, Ganesh Nair, Brandon Doty, Devi G. Nair

## Abstract

**Background:** Iatrogenic atrial septal defect (iASD) is a known consequence of transseptal catheterization. Left atrial intracardiac echocardiography (LA ICE) requires additional septal instrumentation, yet data on persistent iASD after pulsed field ablation (PFA) with an LA ICE workflow remain limited. We evaluated the incidence, predictors, and one-year clinical significance of persistent iASD in this setting.

**Methods:** Consecutive patients undergoing PFA for atrial fibrillation with LA ICE were prospectively evaluated with transthoracic echocardiography before ablation and at one year, including systematic agitated saline contrast. Persistent iASD was defined as residual interatrial shunting on color Doppler at follow-up, classified as small (<3 mm), moderate (3–5 mm), or large (>5 mm). Groups were compared by t-test and chi-square test.

**Results:** Among 850 patients, persistent iASD was identified in 153 (18.0%) at one year; 97 (63.4%) were small and 56 (36.6%) moderate, with no large defects. All shunts were left-to-right. No stroke or transient ischemic attack, paradoxical embolism, hypoxemia, right-heart enlargement, or septal closure occurred. Persistent iASD was associated with female sex (64.7% vs 48.1%), longer septal dwell time (52±12 vs 31±11 min), higher left atrial pressure (28±4 vs 12±3 mmHg), lower LVEF (32±11% vs 54±14%), and larger-caliber sheaths (≥17 Fr; 80.4% vs 48.2%; all p<0.001).

**Conclusions:** Persistent iASD following PFA with LA ICE occurs in approximately one in five patients but is predominantly small, exclusively left-to-right, and clinically benign at one year. Persistence is associated with mechanical and hemodynamic factors, particularly sheath caliber, rather than the ablation energy source.

**Nonstandard Abbreviations and Acronyms:** AF, atrial fibrillation; iASD, iatrogenic atrial septal defect; ICE, intracardiac echocardiography; LA, left atrial; LVEF, left ventricular ejection fraction; OD, outer diameter; PFA, pulsed field ablation; RF, radiofrequency; TIA, transient ischemic attack.

**Data Availability:** The data that support the findings of this study are available from the corresponding author upon reasonable request.

**Central Illustration:** **Central Illustration.**
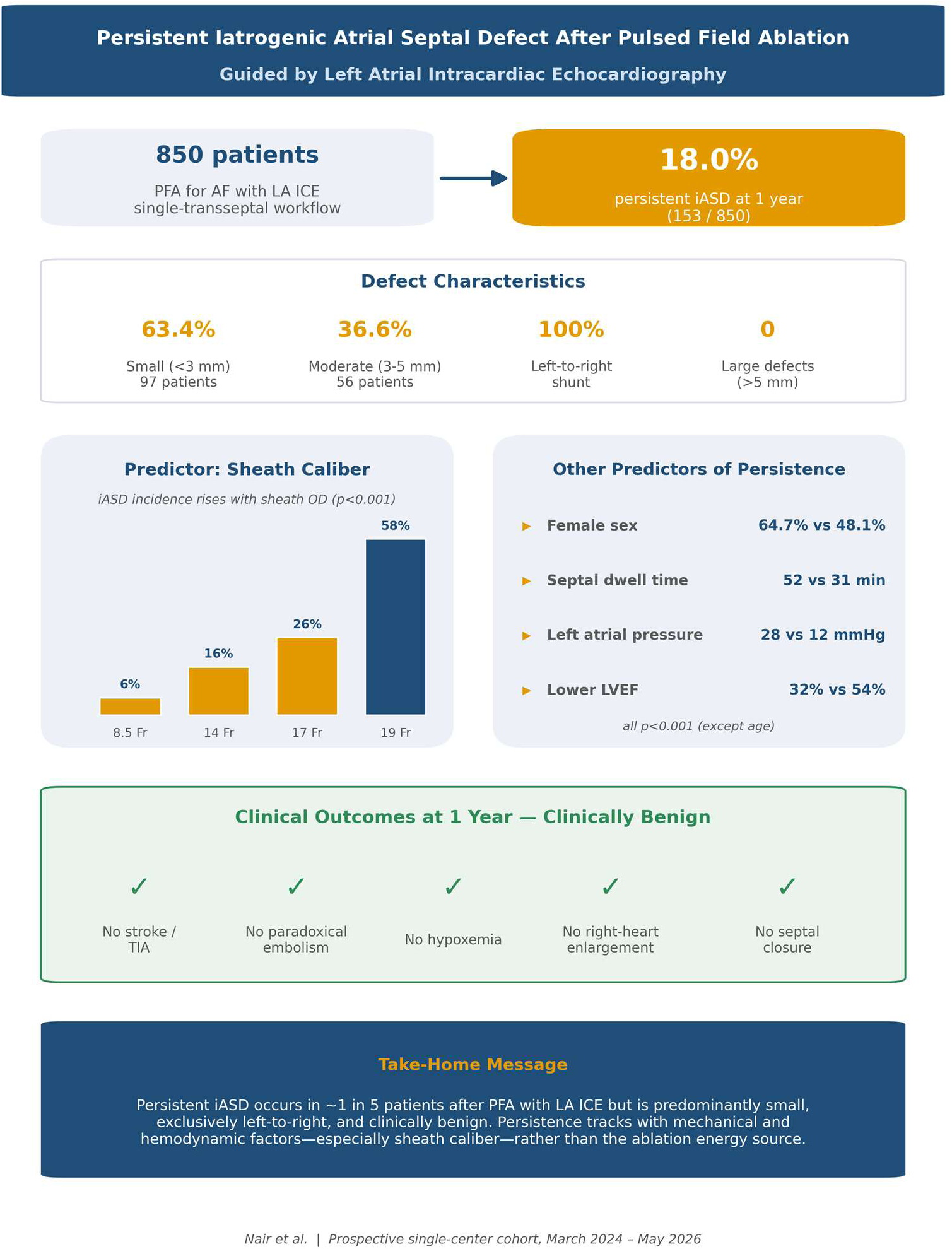
Persistent iASD occurred in 18.0% of 850 patients undergoing PFA with LA ICE but was predominantly small, exclusively left-to-right, and clinically benign at one year. Persistence tracked with mechanical and hemodynamic factors, most notably transseptal sheath caliber, rather than the ablation energy source.

## Introduction

Catheter ablation is an established therapy for symptomatic atrial fibrillation (AF) and is now recommended as first-line rhythm-control therapy in selected patients under contemporary guidelines.^1^ Pulsed field ablation (PFA), a nonthermal modality based on irreversible electroporation, has emerged as a preferred energy source because of its preferential myocardial selectivity and reduced risk of collateral injury compared with thermal techniques.

Regardless of energy source, access to the left atrium requires transseptal puncture, which carries the risk of iatrogenic atrial septal defect (iASD). Transient interatrial communication is common immediately after transseptal catheterization and usually resolves spontaneously; however, a proportion of defects persist. Prior series using radiofrequency and cryoballoon ablation have reported persistent iASD in approximately 10% to 25% of patients, with most defects small and clinically silent, although under-recognition has been described.^1–6^

The clinical relevance of these findings is heightened by the increasing adoption of left atrial intracardiac echocardiography (LA ICE), in which the imaging catheter is advanced across the interatrial septum into the left atrium. This approach adds instrumentation at the septum and may increase septal stretch, prolong dwell time, and alter healing dynamics. Whether these factors translate into a higher rate of persistent iASD in the contemporary PFA era is not well defined, as most existing data predate both PFA and routine LA ICE.

We therefore sought to evaluate the incidence, predictors, and one-year clinical outcomes of persistent iASD in a large prospective cohort of patients undergoing PFA with an LA ICE–guided, single-transseptal workflow.

## Methods

### Study design and population

This was a prospective, non-randomized, single-center imaging cohort study of consecutive patients with atrial fibrillation undergoing pulsed field ablation (PFA) at St. Bernards Medical Center between March 2024 and April 2025, with final follow-up completed in May 2026. All patients were treated using a workflow incorporating left atrial intracardiac echocardiography (LA ICE). Patients without complete baseline and one-year follow-up imaging were excluded from the final analysis, as were cases performed using four-dimensional (4D) ICE and those requiring intraprocedural sheath exchange; all included procedures used a zero-exchange transseptal approach. The study was conducted in accordance with the Declaration of Helsinki and approved by the St. Bernards Institutional Review Board (protocol IASD2024-0412); the requirement for informed consent was waived given the observational nature of the study. None of the patients reported here have been included in any previous publication.

### Ablation procedure

All procedures were performed using a single, zero-exchange transseptal puncture technique under standard procedural conditions. Transseptal puncture was performed under radiofrequency (RF) wire guidance in all cases. Following transseptal access, a commercially available transseptal or access sheath was advanced into the left atrium. Ablation was performed using one of several PFA systems approved for clinical use in the United States, each with its corresponding transseptal or delivery sheath: the Sphere-9 system (Medtronic, Minneapolis, MN) with an Agilis 8.5 Fr steerable sheath (Abbott, St. Paul, MN); the VARIPULSE system (Biosense Webster, Irvine, CA) with the VIZIGO sheath; the PULSESELECT system (Medtronic) with the FlexCath Contour sheath; the FARAWAVE system (Boston Scientific, Marlborough, MA) with the FARADRIVE sheath or an Agilis 13 Fr sheath; and the Globe Pulsed Field Ablation System (Kardium, Burnaby, BC, Canada) with the Globe sheath. All sheath dimensions are reported as outer diameter (OD), ranging from 8.5 to 19 Fr, with the 19 Fr Globe sheath used in a limited number of cases. The Volt PFA system, which employs the same Agilis 13 Fr sheath, was approved later and was not included in this cohort. A 9 Fr ViewFlex Xtra ICE catheter (Abbott, St. Paul, MN), used in all cases, was then advanced across the interatrial septum to enable imaging from within the left atrium. PFA was performed according to standard institutional protocols, with pulmonary vein isolation confirmed by established electrophysiologic criteria. Procedural variables collected prospectively included transseptal sheath size, left atrial septal dwell time (defined as the interval from initial transseptal access to final withdrawal of all left atrial catheters), and invasively measured left atrial pressure.

### Echocardiographic assessment

Transthoracic echocardiography was performed before ablation and again at approximately 12 months. Iatrogenic atrial septal defect (iASD) was defined as residual interatrial shunting identified by color Doppler imaging at the 12-month follow-up study, and persistent iASD as a defect present at one year. Color Doppler imaging was used for both defect identification and sizing, with defect size measured at the maximal width of the residual interatrial communication and categorized as small (<3 mm), moderate (3–5 mm), or large (>5 mm). Agitated saline (bubble) contrast studies were systematically performed in all patients as an adjunctive technique; given their increased sensitivity for detecting small residual interatrial communications, they were used to confirm the presence and direction of shunting but not for primary defect sizing. Transesophageal echocardiography was not used in any subset. Right atrial and right ventricular size were assessed qualitatively using standard echocardiographic criteria. All studies were interpreted by experienced readers blinded to procedural characteristics and clinical outcomes.

### Clinical outcomes

An elective one-year follow-up visit was arranged before discharge and included clinical interview, physical examination, and echocardiography. The primary endpoint was the presence of persistent iASD at one year. Secondary endpoints were defect size, shunt direction, and clinical events potentially related to interatrial shunting, including stroke or transient ischemic attack, hypoxemia, right-sided chamber enlargement, migraine symptoms, and need for septal closure.

### Statistical analysis

Continuous variables are reported as mean ± standard deviation and categorical variables as counts and percentages. Patients with and without persistent iASD were compared using the Student t-test for continuous variables and the chi-square test for categorical variables. A two-sided p-value <0.05 was considered statistically significant. Analyses were performed using SPSS Statistics version 29.0 (IBM Corp., Armonk, NY, USA).

## Results

### Study population

A total of 850 consecutive patients underwent PFA with LA ICE guidance and had complete baseline and one-year imaging. Transseptal access was obtained with commercially available sheaths ranging from 8.5 to 19 Fr in outer diameter, and intracardiac echocardiography was performed using the ViewFlex Xtra Intracardiac Echocardiography (ICE) Catheter (Abbott, St. Paul, MN) in all cases. Baseline demographic, clinical, and procedural characteristics are summarized in Table 1.

**Table 1.** Baseline and Procedural Characteristics. Continuous variables are mean ± SD; categorical variables are counts/percentages. p-values from Student t-test or chi-square test.

| Variable | Persistent iASD (n=153) | No iASD (n=697) | p-value |
| --- | --- | --- | --- |
| Age (years) | 69 ± 9 | 71 ± 11 | 0.017 |
| Female sex, n (%) | 99 (64.7%) | 335 (48.1%) | <0.001 |
| LVEF (%) | 32 ± 11 | 54 ± 14 | <0.001 |
| Left atrial pressure (mmHg) | 28 ± 4 | 12 ± 3 | <0.001 |
| LA septal dwell time (min) | 52 ± 12 | 31 ± 11 | <0.001 |
| Transseptal sheath OD, n (%) |  |  | <0.001 |
| 8.5 Fr | 18 (11.8%) | 297 (42.6%) |  |
| 14 Fr | 12 (7.8%) | 64 (9.2%) |  |
| 17 Fr | 112 (73.2%) | 328 (47.1%) |  |
| 19 Fr | 11 (7.2%) | 8 (1.1%) |  |
| ICE catheter (ViewFlex Xtra), Fr | 9 | 9 | , |

### Incidence and characteristics of persistent iASD

At one-year follow-up, persistent iASD was identified in 153 patients (18.0%); the remaining 697 (82.0%) had no residual interatrial shunting. Among defects, 97 (63.4% of iASD; 11.4% of the total cohort) were small (<3 mm) and 56 (36.6% of iASD; 6.6% of the cohort) were moderate (3–5 mm). No large defects (>5 mm) were observed. All persistent defects demonstrated left-to-right shunting; no right-to-left or bidirectional shunts were detected (Table 2; Figures 1 and 3).

**Figure 1.**
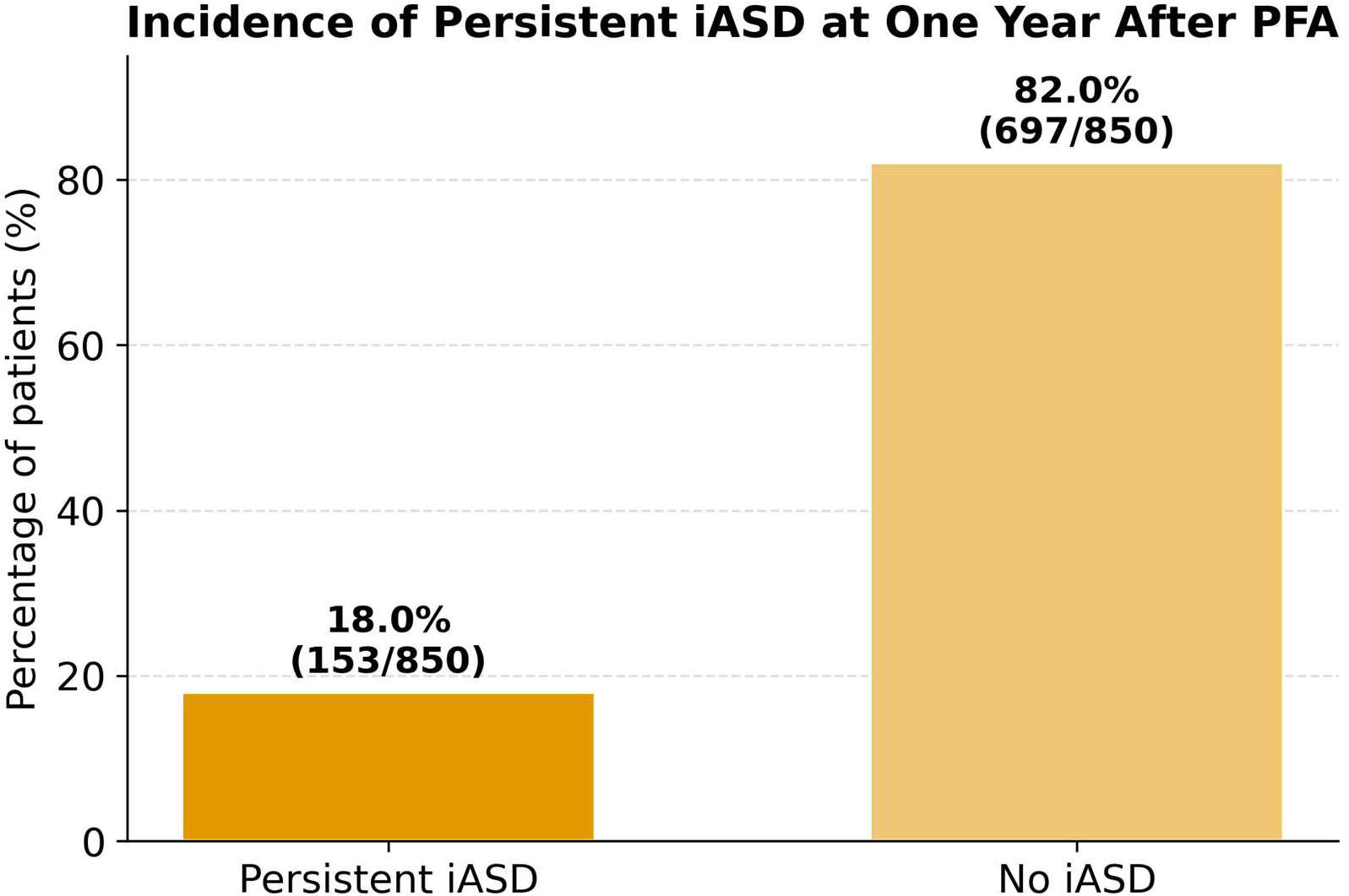
Incidence of persistent iatrogenic atrial septal defect (iASD) following pulsed field ablation with left atrial intracardiac echocardiography. Persistent iASD was identified in 18.0% of patients at one year, with no residual interatrial shunting in 82.0%.

**Figure 2.**
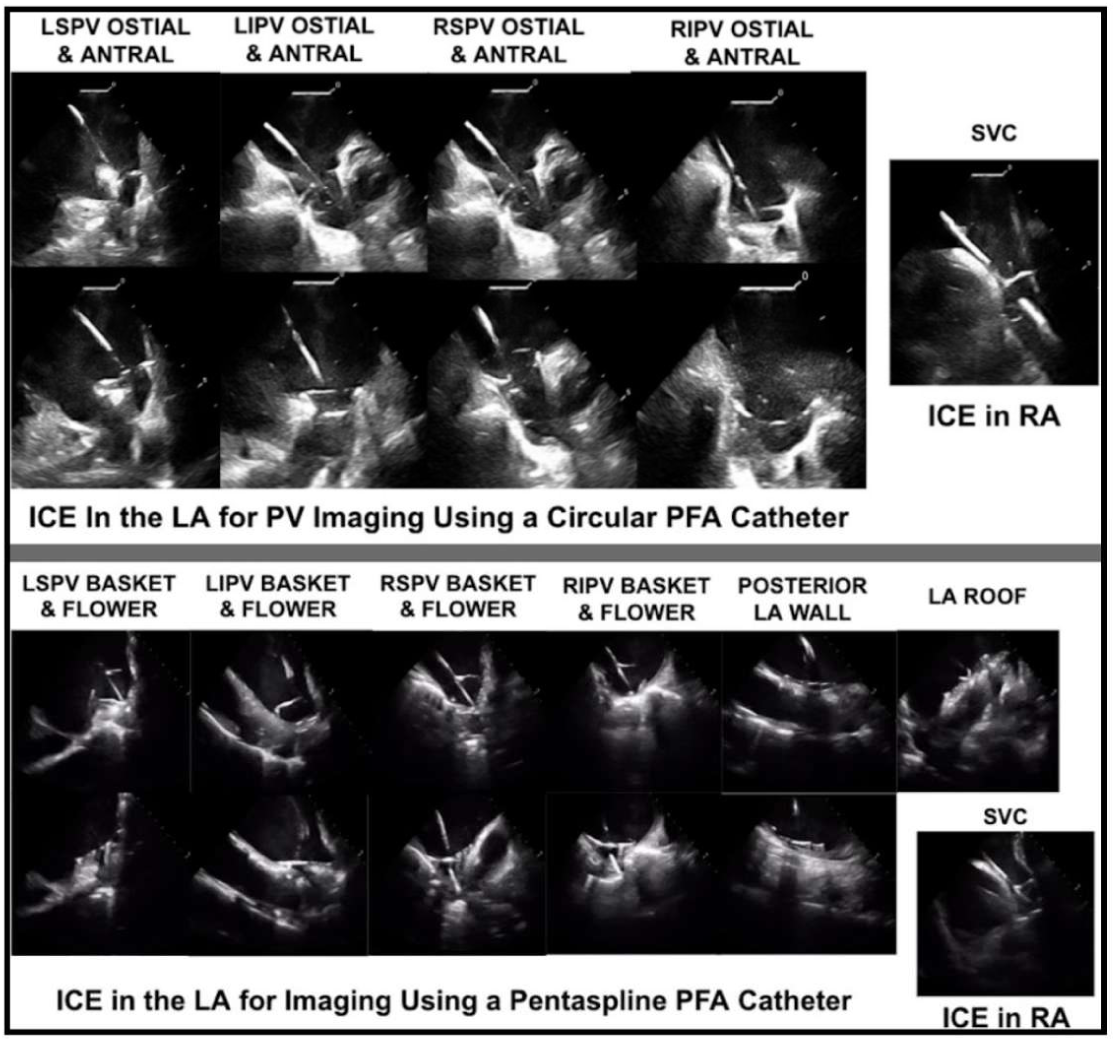
Representative left atrial intracardiac echocardiography imaging during pulsed field ablation. Top panel: pulmonary vein imaging using a circular PFA catheter (LSPV, LIPV, RSPV, and RIPV ostial and antral views; SVC and ICE-in-RA reference views). Bottom panel: imaging using a pentaspline PFA catheter (basket and flower configurations at each pulmonary vein, posterior left atrial wall, and left atrial roof).

**Figure 3.**
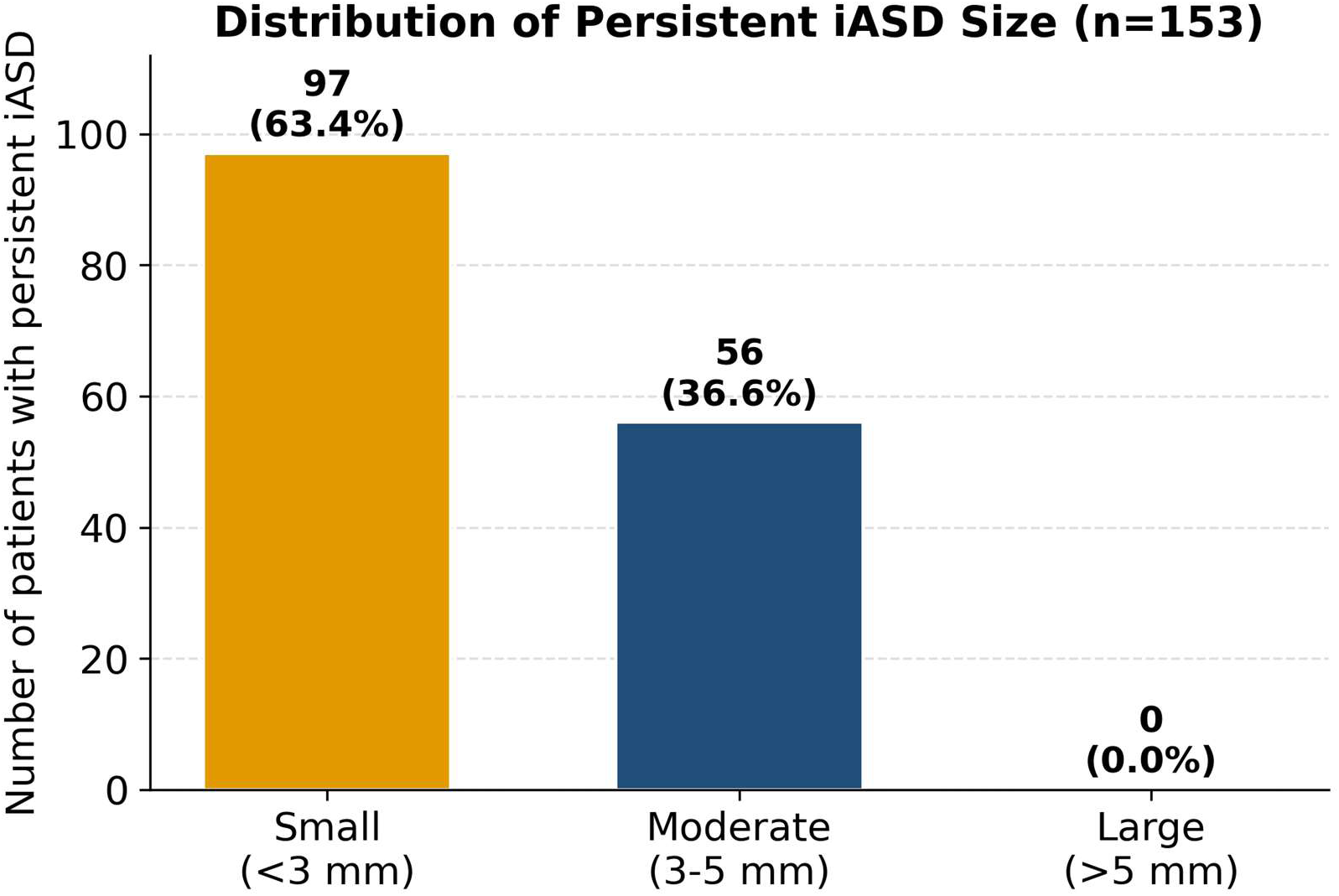
Distribution of persistent iASD size at one year. Of 153 persistent defects, 97 (63.4%) were small (<3 mm) and 56 (36.6%) were moderate (3–5 mm); no large defects (>5 mm) were observed.

**Table 2.** Incidence and Characteristics of Persistent iASD at One Year.

| Variable | Value |
| --- | --- |
| Total cohort | 850 |
| Persistent iASD | 153 (18.0%) |
| No persistent iASD | 697 (82.0%) |
| Small iASD (<3 mm) | 97 (11.4% of cohort; 63.4% of iASD) |
| Moderate iASD (3–5 mm) | 56 (6.6% of cohort; 36.6% of iASD) |
| Large iASD (>5 mm) | 0 |
| Left-to-right shunt | 153 (100%) |
| Right-to-left / bidirectional shunt | 0 |
| Right heart enlargement | 0 |
| Septal closure required | 0 |

### Clinical outcomes at one year

No patient experienced stroke or transient ischemic attack, paradoxical embolism, hypoxemia, or right-sided chamber enlargement, and none required percutaneous or surgical septal closure. Migraine symptoms were reported in 1.2% of patients and resolved by six months in the majority; symptoms persisted to one year in only four patients (Table 3; Figure 4).

**Figure 4.**
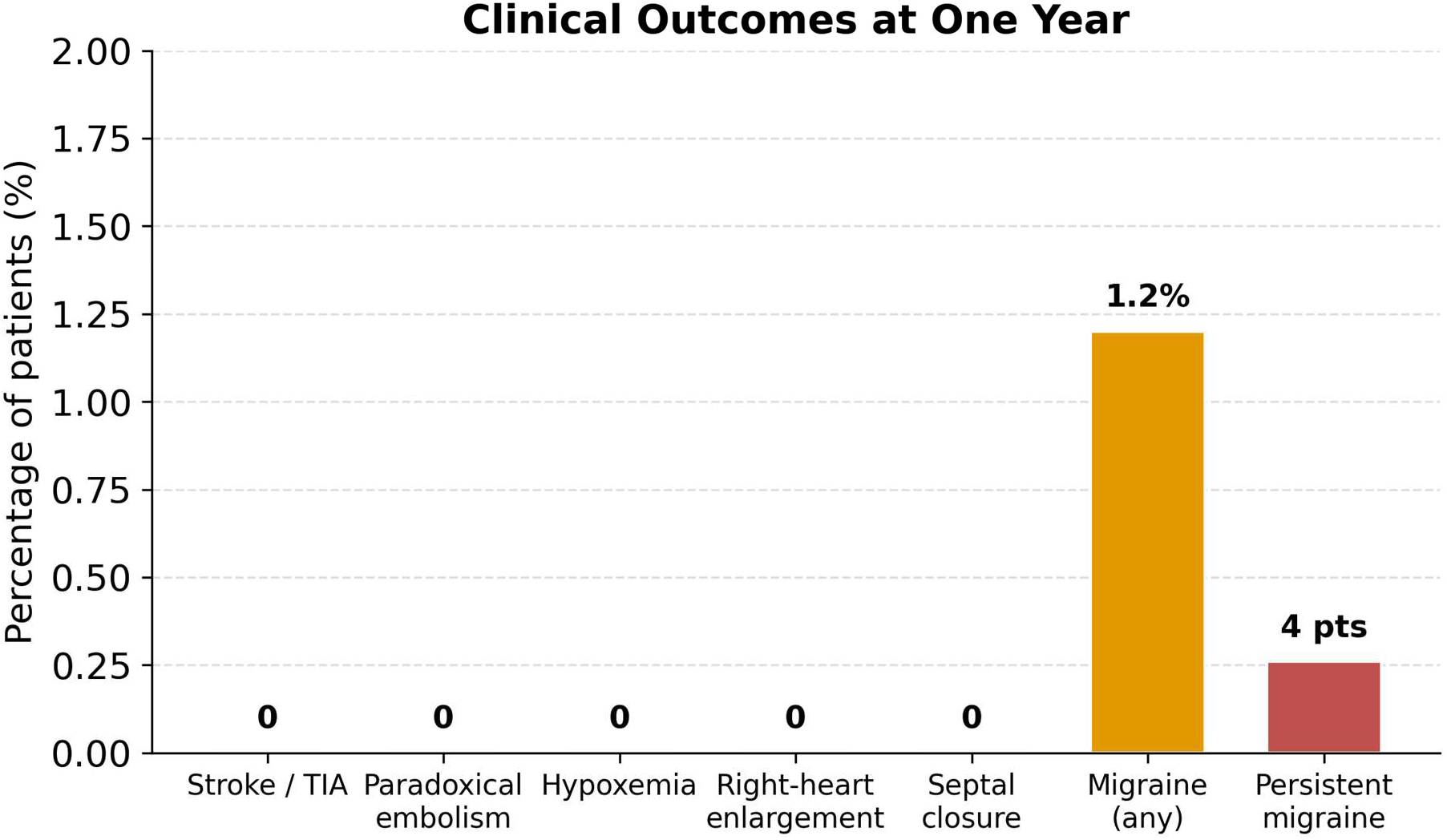
Clinical outcomes associated with persistent iASD at one year. No patient experienced stroke or transient ischemic attack, paradoxical embolism, hypoxemia, right-sided chamber enlargement, or required septal closure; migraine symptoms occurred in 1.2% and persisted in only four patients.

**Table 3.** Clinical Outcomes at One Year.

| Outcome | Value |
| --- | --- |
| Stroke / transient ischemic attack | 0 |
| Paradoxical embolism | 0 |
| Hypoxemia | 0 |
| Right-sided chamber enlargement | 0 |
| Migraine symptoms | 1.2% |
| Persistent migraine at 1 year | 4 patients |
| Septal closure | 0 |

### Predictors of persistent iASD

Compared with patients without persistent iASD, those with persistent iASD were more frequently female (64.7% vs 48.1%, p<0.001) and slightly younger (69±9 vs 71±11 years, p=0.017). Patients with persistent iASD also demonstrated significantly longer left atrial septal dwell time (52±12 vs 31±11 minutes, p<0.001), higher mean left atrial pressure (28±4 vs 12±3 mmHg, p<0.001), and lower left ventricular ejection fraction (32±11% vs 54±14%, p<0.001). The distribution of transseptal sheath outer diameter also differed significantly between groups (p<0.001), with a stepwise increase in iASD incidence across increasing sheath caliber: 5.7% (18/315) with 8.5 Fr, 15.8% (12/76) with 14 Fr, 25.5% (112/440) with 17 Fr, and 57.9% (11/19) with 19 Fr sheaths. Larger-diameter sheaths (≥17 Fr) were used in 80.4% of patients with persistent iASD compared with 48.2% of those without (Table 1; Figure 5).

**Figure 5.**
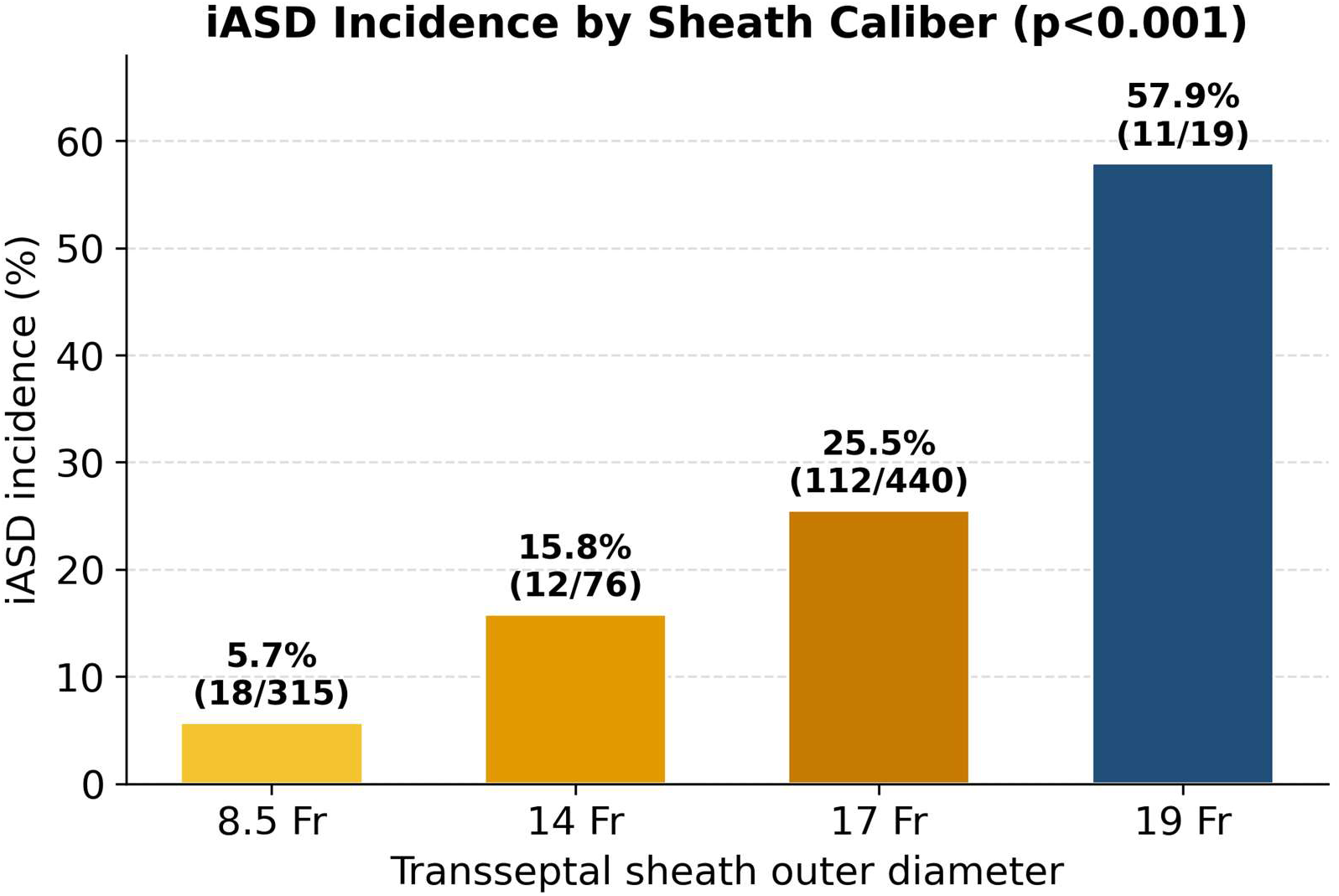
Incidence of persistent iASD by transseptal sheath outer diameter, demonstrating a stepwise increase with larger sheath caliber (5.7% at 8.5 Fr to 57.9% at 19 Fr; p<0.001). Because each PFA system uses a sheath of characteristic caliber, sheath outer diameter is closely linked to the ablation platform employed.

## Discussion

In this prospective cohort of 850 patients undergoing pulsed field ablation (PFA) with left atrial intracardiac echocardiography (LA ICE), persistent iatrogenic atrial septal defect (iASD) was present in 18.0% at one year. Four principal findings emerge. First, persistent iASD is not uncommon following PFA with LA ICE and does not uniformly close within the first postprocedural year. Second, despite this frequency, the overwhelming majority of defects were small, exclusively left-to-right, and produced no measurable clinical consequence. Third, persistence was associated with mechanical and hemodynamic factors, longer septal dwell time, higher left atrial pressure, larger sheath caliber, and reduced left ventricular systolic function, rather than the ablation energy source. Fourth, to our knowledge this represents one of the largest cohorts with structured one-year septal surveillance in the contemporary PFA era incorporating an LA ICE workflow.

### Incidence in context

Prior series using radiofrequency or cryoballoon ablation have reported persistent iASD in roughly 10% to 25% of patients, with most defects small and clinically silent.³⁻⁶ Our incidence of 18.0% falls within this range, indicating that PFA combined with LA ICE does not appear to increase the risk of septal persistence relative to conventional thermal modalities. This is notable because most earlier studies did not incorporate routine LA ICE, which requires additional catheter manipulation across the septum and might be expected to prolong instrumentation and impair healing. Our findings suggest that, despite these theoretical concerns, the septal consequences of an LA ICE workflow remain modest and clinically benign. Notably, our incidence of 18.0% closely mirrors the 18.1% reported in the prospective EVITA study of cryoballoon and radiofrequency ablation (n=94),⁵ and our identification of larger transseptal sheath caliber as a predictor of persistence is concordant with, and extends the recent observations of Kamada et al.,⁶ now in a markedly larger cohort (n=850) and in the contemporary PFA era incorporating LA ICE.

### Mechanisms of persistent iASD

The formation and persistence of iASD following transseptal catheterization are primarily mechanical. Prolonged septal dwell time, larger-diameter sheaths, and elevated left atrial pressure likely promote septal stretch and impair tissue coaptation, while a sustained left-to-right pressure gradient may keep a defect patent and limit spontaneous closure. The association between reduced left ventricular ejection fraction and persistent iASD is consistent with this framework, plausibly reflecting elevated left-sided filling pressures and altered atrial compliance. The higher prevalence among female patients may relate to differences in septal thickness or tissue characteristics, though this observation is hypothesis-generating and warrants dedicated study. Importantly, given the nonthermal mechanism of PFA and the typical location of energy applications away from the septum, it is unlikely that energy delivery itself contributes materially to septal injury, reinforcing that septal outcomes are determined by access technique and mechanical factors rather than ablation modality.

### PFA system versus sheath caliber

A notable feature of our cohort is that sheath outer diameter is closely linked to the specific PFA system employed, because each platform uses a delivery or transseptal sheath of characteristic caliber, for example, the Sphere-9 system (Agilis 8.5 Fr) at the smaller end and the Globe system (19 Fr) at the larger end. As a result, sheath caliber and PFA system are collinear, and their independent contributions cannot be fully disentangled in this observational dataset. Several considerations nonetheless favor sheath caliber, a mechanical factor, as the dominant driver. First, iASD incidence rose monotonically with increasing sheath outer diameter across systems, from 5.7% with 8.5 Fr sheaths to 57.9% with 19 Fr sheaths. Second, two distinct platforms, the Sphere-9 and VARIPULSE systems, share the same 8.5 Fr sheath, and both fell within the lowest-incidence stratum, indicating that the access caliber rather than the energy platform governs septal persistence. Third, all included systems deliver nonthermal energy and target tissue away from the interatrial septum, making a system-specific biological effect on septal healing unlikely. Fourth, the procedural and hemodynamic factors associated with persistence in prior thermal-ablation cohorts, dwell time, left atrial pressure, and access caliber, were operative here as well. Taken together, these observations suggest that the apparent system-level differences are most plausibly explained by the caliber of the transseptal instrumentation rather than by the energy platform itself.

### Clinical implications

The clinical significance of persistent iASD after AF ablation has been debated. In our cohort, no patient developed right-heart volume overload, hypoxemia, or paradoxical embolism, and none required septal closure at one year. These data suggest that routine surveillance or intervention for small, asymptomatic iASD following PFA is unlikely to be necessary. Nevertheless, recognizing the factors associated with persistence may inform procedural optimization: minimizing septal dwell time, limiting sheath caliber where feasible, and attending to periprocedural hemodynamics are reasonable strategies. Selective follow-up imaging may be considered in higher-risk patients, those with elevated left atrial pressure or reduced ventricular function, identified in this analysis.

### Limitations

This study has several limitations. First, it was a single-center observational study, which may limit generalizability. Second, although agitated saline contrast was performed systematically in all patients to enhance detection of small shunts, assessment relied on transthoracic echocardiography, which has lower spatial resolution than transesophageal imaging for very small defects. Third, imaging was obtained at one year, so the temporal evolution of defects between the procedure and follow-up was not systematically characterized. Fourth, although procedural and clinical associations were identified, the observational design precludes inference of causality. Fifth, because each PFA system was used with a sheath of characteristic caliber, PFA system and sheath outer diameter were collinear, and our analysis cannot definitively separate their respective contributions to septal persistence. Finally, outcomes beyond one year remain unknown, and multicenter validation is warranted.

## Conclusion

In a large prospective cohort undergoing PFA with LA ICE guidance, persistent iASD was present in 18.0% of patients at one year but was predominantly small, exclusively left-to-right, and not associated with adverse clinical outcomes. Persistence appears to be driven by procedural and hemodynamic factors rather than the ablation energy source. These findings support the overall safety of contemporary PFA workflows incorporating LA ICE while identifying modifiable factors, septal dwell time, sheath caliber, and hemodynamic status, that may further optimize transseptal technique and inform follow-up strategies.

## Novelty and Significance

### What Is Known

- Transseptal puncture for left atrial access can create an iatrogenic atrial septal defect (iASD) that persists in roughly 10–25% of patients after radiofrequency or cryoballoon ablation.
- Most persistent iASDs are small and clinically silent, but data in the contemporary pulsed field ablation (PFA) era, particularly with left atrial intracardiac echocardiography (LA ICE), which adds instrumentation across the septum, are limited.

### What New Information Does This Article Contribute?

- In 850 patients undergoing PFA with an LA ICE–guided, single-transseptal workflow, persistent iASD occurred in 18.0% at one year but was predominantly small, exclusively left-to-right, and clinically benign, with no stroke, paradoxical embolism, hypoxemia, right-heart enlargement, or need for septal closure.
- iASD incidence increased stepwise with transseptal sheath outer diameter (5.7% to 57.9%), and two different PFA systems sharing the smallest-caliber sheath had the same low incidence, indicating that sheath caliber, not the ablation energy source, drives septal persistence.
- PFA with LA ICE does not increase the risk of persistent iASD relative to thermal ablation, supporting the safety of contemporary workflows while identifying modifiable procedural factors.

In this prospective cohort of 850 patients undergoing pulsed field ablation for atrial fibrillation with a left atrial intracardiac echocardiography (LA ICE)–guided, single-transseptal workflow, persistent iatrogenic atrial septal defect was present in 18.0% at one year. Defects were predominantly small and exclusively left-to-right, and none produced clinically meaningful sequelae. Persistence was associated with mechanical and hemodynamic factors, most prominently transseptal sheath caliber: iASD incidence rose stepwise with sheath outer diameter, and two different PFA platforms that share the smallest-caliber sheath had the same low incidence, indicating that access caliber rather than the energy source governs septal persistence. These data, from one of the largest contemporary cohorts with structured one-year septal surveillance, indicate that PFA with LA ICE does not increase the risk of persistent iASD relative to thermal ablation, and they identify modifiable procedural targets, sheath caliber, septal dwell time, and hemodynamics, to optimize transseptal technique.

## IRB approval

St. Bernards Institutional Review Board, protocol IASD2024-0412; requirement for informed consent waived (observational study).

